# Love during lockdown: findings from an online survey examining the impact of COVID-19 on the sexual practices of people living in Australia

**DOI:** 10.1101/2020.08.10.20171348

**Authors:** Jacqueline Coombe, Fabian Kong, Helen Bittleston, Hennie Williams, Jane Tomnay, Alaina Vaisey, Sue Malta, Jane Goller, Meredith Temple-Smith, Louise Bourchier, Andrew Lau, Eric P.F. Chow, Jane S Hocking

**Affiliations:** Melbourne School of Population and Global Health, The University of Melbourne, 207 Bouverie Street, Carlton, VIC 3053, Australia; Melbourne Sexual Health Centre, Alfred Health, 580 Swanston Street, Carlton, VIC 3053, Australia; Centre for Excellence in Rural Sexual Health, Department of Rural Health, University of Melbourne VIC 3010; National Ageing Research Institute, Poplar Road, Parkville, VIC 3052; Department of General Practice, The University of Melbourne, 780 Elizabeth Street, Carlton 3010, Australia; Central Clinical School, Monash University, 99 Commercial Road, Melbourne, VIC 3004, Australia

**Author notes:** Corresponding author: Jacqueline Coombe, Melbourne School of Population and Global Health, The University of Melbourne, 207 Bouverie Street, Carlton, 3053, Australia. E.

## Abstract

**Introduction:** Australia recorded its first case of COVID-19 in late January 2020. On 22^nd^ March 2020, amid increasing daily case numbers, the Australian Government implemented lockdown restrictions to help ‘flatten the curve’. Our study aimed to understand the impact of lockdown restrictions on sexual and reproductive health. Here we focus on sexual practices.

**Methods:** An online survey was open from the 23^rd^ April 2020 to the 11^th^ May 2020. Participants were recruited online via social media and other networks and were asked to report on their sexual practices in 2019 and during lockdown. Logistic regression was used to calculate the difference (including 95% confidence intervals) in the proportion of sex practices between time periods.

**Results:** Of the 1187 who commenced the survey, 965 (81.3%) completed it. Overall 70% were female and 66.3% were aged 18 to 29 years. Most (53.5%) reported less sex during lockdown than in 2019. Compared with 2019, participants were more likely to report sex with a spouse (35.3% vs 41.7%; difference=6.4%; 95%CI: 3.6, 9.2) and less likely to report sex with a girl/boyfriend (45.1% vs 41.8%; diff=-3.3%; 95%CI: -7.0, -0.4) or with casual hook-up (31.4% vs 7.8%; 95%CI:-26.9, -19.8). Solo sex activities increased, 14.6% (123/840) reported using sex toys more often and 26.0% (218/838) reported masturbating more often. Dating app use decreased during lockdown compared with 2019 (42.1% vs 27.3%; difference= -14.8%; 95%CI: -17.6, -11.9). Using dating apps for chatting/texting (89.8% vs 94.5%; diff=4.7%; 95%CI:1.0, 8.5) and for setting up virtual dates (2.6% vs 17.2%; diff=14.6%; 95%CI:10.1, 19.2) increased during lockdown.

**Conclusion:** Although significant declines in sexual activity during lockdown were reported, people did not completely stop engaging in sexual activities during the pandemic, highlighting the importance of ensuring availability of normal sexual and reproductive health services during global emergencies.

**KEY MESSAGES:**

- Sexual activity declined among our participants during the COVID-19 lockdown restrictions in Australia, with more than half reporting having less sex than in 2019
- Sexual practices also changed during lockdown, with more people reporting solo sex activities like masturbating alone or using a sex toy.
- Use of dating apps also declined among our participants. Of those still using apps, we saw increased use for chatting/texting and setting up virtual dates.

## INTRODUCTION

In late January 2020, Australia recorded its first case of COVID-19[1]. In response to rapidly increasingly daily case numbers[1], the Australian Government began to introduce several measures in an attempt to ‘flatten the curve’. On the 22^nd^ March, Stage 1 restrictions were announced, including the temporary closure of non-essential businesses and services, limiting the size of non-essential gatherings, promoting social distancing and advising against non-essential travel[2]. As daily case numbers continued to top 300 in the following week[1], strict lockdown measures were implemented from the 29^th^ March, with people asked to remain in their homes and only leave for four activities: shopping for essential goods and services, to exercise, to seek medical care or to attend work or education where these activities could not take place at home[3]. Gatherings were limited to two persons only[4]. Children were home-schooled by their caregivers, and large swathes of the workforce began working from home indefinitely. In addition to the closure of the international border, most state and territory borders were also closed, effectively preventing interstate travel (and at the time of writing, have not yet reopened). These strict lockdown measures continued until the 8^th^ May 2020, when the Australian Government announced a plan for the easing of restrictions and a ‘COVID-safe Australia’[5] as case numbers across the country consistently declined to below 30^1^ a day[1].

During this isolation period, colloquially referred to by the Australian public as ‘iso’, most people significantly reduced their activities outside their homes[6], and a survey conducted by the Australian Bureau of Statistics found 94% of participants were keeping their distance from those outside their household[7]. Intimate relationships were not exempt from the impact of the lockdown. As restrictions were enforced across the country, confusion abounded among those with non-cohabitating intimate partners, as they wondered whether visits were allowed under the new restrictions. While some states banned people, including non-cohabitating intimate partners, to meet unless they were exercising together or providing care[8], others changed course during lockdown and ultimately made an exception to allow non-cohabitating partners to meet[9, 10]. For those isolating with their partners, some prophesised that Australia is much more likely to experience a declining birth rate than a baby-boom nine months post-lockdown[11] as people grapple with the social, economic and health impacts of the pandemic. For those without regular partners, sexual health organisations and state health departments advised against casual sex during lockdown but provided advice on how to still enjoy sexual pleasure in the absence of physically present partner/s [12-14]. Dating apps like Tinder and Hinge included in-app safety messages about how to connect with new partners while maintaining social distancing, suggesting using video platforms to set up virtual dates[15].

Evidently, the COVID-19 pandemic and the lockdown it prompted are likely to have an impact on the sexual practices and sexual health of people living in Australia. We implemented a serial cross-sectional survey that aimed to investigate the impact of COVID-19 on sexual and reproductive health of people living in Australia. In this paper we report on the results from the first survey and explore the impact of lockdown on people’s sexual practices.

## METHODS

### Sexual and Reproductive Health during COVID-19 Online Survey

The first survey was administered online between the 23^rd^ April to 11^th^ May. All Australian states and territories were under lockdown to varying degrees during this period, with restrictions easing from the 8^th^ May. The survey comprised questions pertaining to the impact of COVID-19 on sexual practices and sexual and reproductive health. As a repeated cross-sectional survey, participants were asked to provide their month and year of birth and their ‘porn star name’ (name of their first pet and the name of the first street they lived on) to enable participant tracking over time[16]. Participants were also asked to provide an email address if they wished to be contacted for future surveys. Email addresses were removed from the dataset and not used to link responses. Repeat waves of the survey will be administered every six to eight weeks across the remainder of 2020, and a cohort analysis will be undertaken on those participants who respond more than once. In this paper, we report on data pertaining to the baseline survey only (subsequent waves of data collection are ongoing). This study was approved by the University of Melbourne Human Research Ethics Committee (ID: 2056693).

### Participants

Participants were recruited via various means, including the research team’s existing networks (for example, emailing the recruitment flyer to colleagues and sexual and reproductive health newsletter lists for distribution, posting on student noticeboards and personal social media accounts and posting the recruitment flyer on our research group’s Twitter account) and via paid Facebook ads. Participants were asked to pass the link to the survey on to their own networks. People were eligible to participate if they were aged 18 years or older and living in Australia at the time of the survey. Participants clicked on a link that took them to the survey page where they were provided with a plain language statement. If they were interested in participating, they were asked to confirm that: they were aged 18 years or over, they understood what the survey was about, and they consented to participate.

### Data collection

Survey questions included trends and changes in sexual practices, intimate relationships, access to essential goods and services, trends and changes in contraceptive use and pregnancy intentions. Sex was defined as physical contact with other people for sexual pleasure including oral sex and mutual masturbation. For several questions, participants were asked to report on two time periods: their experiences and practices during all of 2019 and during ‘*lockdown*’ (the period after March 22^nd^ when restrictions commenced). Participants were also asked to report on the frequency of sexual activity by commenting on whether their sexual activity was less, the same or more during lockdown than in 2019. They were also asked to report whether specific sexual practices (such as masturbating alone or oral sex) were being performed the same amount, less often, more or stopped completely because of COVID-19. We also collected sociodemographic data.

### Data analysis

A sample size of 800 would allow us to detect a difference in paired proportions of 6% (55% vs 56%) assuming a correlation of 0.25, power of 80% and alpha of 0.05. Descriptive statistics were used to describe the sociodemographic characteristics of participants. Logistic regression was used to determine the difference in proportion between the two time periods (lockdown minus 2019) for categorical variables adjusting for clustering at the participant level. The difference in the proportion and its corresponding 95% confidence intervals are reported. We fitted interaction terms between the time period (lockdown versus 2019) and the variables of gender, age, sexuality and relationship status in each logistic regression model to examine whether the difference in reported activities between time periods varied across different categories of these variables (for example, did the difference in app use between the two time periods vary between those aged 18 to 29 years and those aged 30+ years). We also used Chi square tests and tests for equality of proportions to investigate associations between categorical variables where indicated and t or Mann Whitney tests to compare continuous variables between two groups. As not everyone completed all questions, missing data are excluded from all analyses, but denominators are provided to put these missing data into context. To assess response bias, we compared the gender, age and sexuality of those who completed and did not complete the survey. All statistical analyses were performed using Stata SE 16.0 for Windows.

## RESULTS

### Demographics

A total of 1187 people consented to participate and commenced the survey; 965 (81.3%) completed it and were included in the analysis. Those who completed the survey were older on average than those who did not complete (28.6 vs 26.2 years, p<0.01) but there was no difference in gender (p=0.44) or sexuality (p=0.11). Overall, 70.0% were female, 66.3% were aged under 30 (median age=24; IQR=20-33; range 18-77), 82.7% reported Australia as their country of birth and 65.7% indicated they were heterosexual. In 2019, 80.2% reported being employed, but this fell to only 63.4% during lockdown. Overall, 9.1% reported testing for COVID-19, but none had tested positive. Sociodemographic data are reported in Table 1.

**Table 1.** Sociodemographic characteristics of survey participants

| Variable | n/N | % |
| --- | --- | --- |
| **Gender\*** |  |  |
| Male | 247/962 | 25.7 |
| Female | 673/962 | 70.0 |
| Gender diverse | 42/962 | 4.4 |
| **Age** |  |  |
| 18-29 | 636/959 | 66.3 |
| 30+ | 323/959 | 33.7 |
| **Relationship Status†** |  |  |
| In a relationship and cohabitating | 346/960 | 36.0 |
| In a relationship but not cohabitating | 248/960 | 25.8 |
| Single | 329/960 | 34.3 |
| Other | 37/960 | 3.4 |
| **Sexual identity ‡** |  |  |
| Heterosexual/straight | 632/962 | 65.7 |
| Men who have sex with men [MSM] | 85/962 | 8.8 |
| Women who have sex with women [WSW] | 197/962 | 20.5 |
| Other | 48/962 | 5.0 |
| **Country of birth** |  |  |
| Australia | 783/946 | 82.7 |
| Other | 163/946 | 17.3 |
| **Aboriginal and/or Torres Strait Islander** |  |  |
| Yes | 26/951 | 2.7 |
| **Highest level of education** |  |  |
| Primary / Secondary school | 254/958 | 26.5 |
| Certificates / Diplomas / Apprenticeships | 144/958 | 15.0 |
| University | 560/958 | 58.5 |
| **Employment in 2019§** |  |  |
| Employed (incl. full time, part time, casual) | 769/959 | 80.2 |
| Student | 326/959 | 34.0 |
| Unemployed and looking for work¶ | 27/959 | 2.8 |
| Other\|\| | 48/959 | 5.0 |
| **Employment in lockdown§** |  |  |
| Employed (incl. full time, part time, casual) | 606/956 | 63.4 |
| Student | 287/956 | 30.0 |
| Stood down due to COVID-19 and/or unemployed looking for work¶ | 135/956 | 14.1 |
| Other\|\| | 58/956 | 6.1 |
| **Tested for COVID-19** |  |  |
| Yes, tested negative | 78/958 | 8.1 |
| Yes, waiting on results | 10/958 | 1.0 |
| No | 870/958 | 90.7 |

N=Number who answered the question, denominator is not always 965 because of missing data; \* Gender diverse includes transgender and non-binary; † 'In a relationship' includes those who selected married/de facto/boyfriend/girlfriend/LAT (living apart together) or long distance relationship; 'cohabitating' defined as living with partner/s; 'Single' includes those who selected single/divorced/widowed (and did not select another relationship); 'Other' includes people who selected polyamorous or multiple partners (and did not select another relationship option, such as single), and those who indicated in comments that they were casually dating, but not exclusive; ‡ MSM includes bisexual and trans men; WSW includes bisexual and trans women; Other includes asexual and non-binary respondents, and those who selected 'something else' ; § Participants can be in multiple categories; || Includes retired, parent/carer, disability support pension, unemployed but not looking for work; ¶ Excluding students

### Sexual activity

Participants reported a median of one sex partner in 2019, (IQR 1-3; range: 0-1000) and a median of one sex partner (IQR 0-1; range: 0-10) during lockdown. Overall, 89.8% (847/943) reported sex in 2019 and 60.3% (555/920) during lockdown (diff=-29.5%; 95%CI: -32.6, - 26.3). Most participants (472/883; 53.5%) reported less sex during lockdown than during 2019 with a small proportion (126/883; 14.3%) reporting that they were having more sex. There was no difference in the frequency of sex by gender, but it varied across age groups (p<0.01), by sexuality (p<0.01) and by relationship status (p<0.01). Across all variables, MSM were the most likely to report less sex than in 2019 (56/80; 70.0%) and those in a cohabitating relationship were the most likely to report the same amount of sex (146/321; 45.5%), or more sex than in 2019 (62/321; 19.3%, Supplementary Table 1).

Participants were more likely to report having sex with a spouse during lockdown compared with 2019 (41.7% vs 35.3%; diff=6.4%; 95%CI:3.6, 9.2) and less likely to report sex with a girl/boyfriend (41.8% vs 45.1%; diff=-3.3%; 95%CI:-7.0, -0.4) or sex with a casual hook-up (7.8% vs 31.4%; diff=-23.4%; 95%CI: -26.9, -19.8, Supplementary Table 2). The difference in proportions between time periods for each partner type did not vary by sexuality or age, but males were more likely to have sex with their spouse during lockdown (diff= 12.4% for males versus 4.6% for females; p=0.04). Singles had significantly less sex with a girl/boyfriend during lockdown compared with those in cohabitating relationships (diff=-26% vs 0.9%; p<0.01). A small number of participants (10/815, 1.2%) reported participating in group sex, swinging or threesomes since lockdown compared with 2019 (133/815, 16.3%; diff=-15.1%; 95%CI: -17.6, -12.5).

### Sexual practices

When asked whether participants had changed their sexual practices because of COVID-19, 14.6% (123/840) reported that they were using sex toys more often on their own and 26.0% (218/838) reported that they were masturbating more. When stratified by frequency of sex during lockdown, those who reported less or no sex during lockdown were more likely to report using sex toys alone (18.3% vs 8.3%, diff=10.0%; 95%CI: 5.5, 14.6) and masturbating alone (35.6% vs 10.3%; diff=25.3%; 95%CI: 20.0, 30.6) compared with those who reported the same amount or more sex since COVID-19 (Figure 1). A total of 98 participants (11.5%) reported buying a sex toy during lockdown and of these, 24.0% indicated that this was their first.

**Figure 1.**
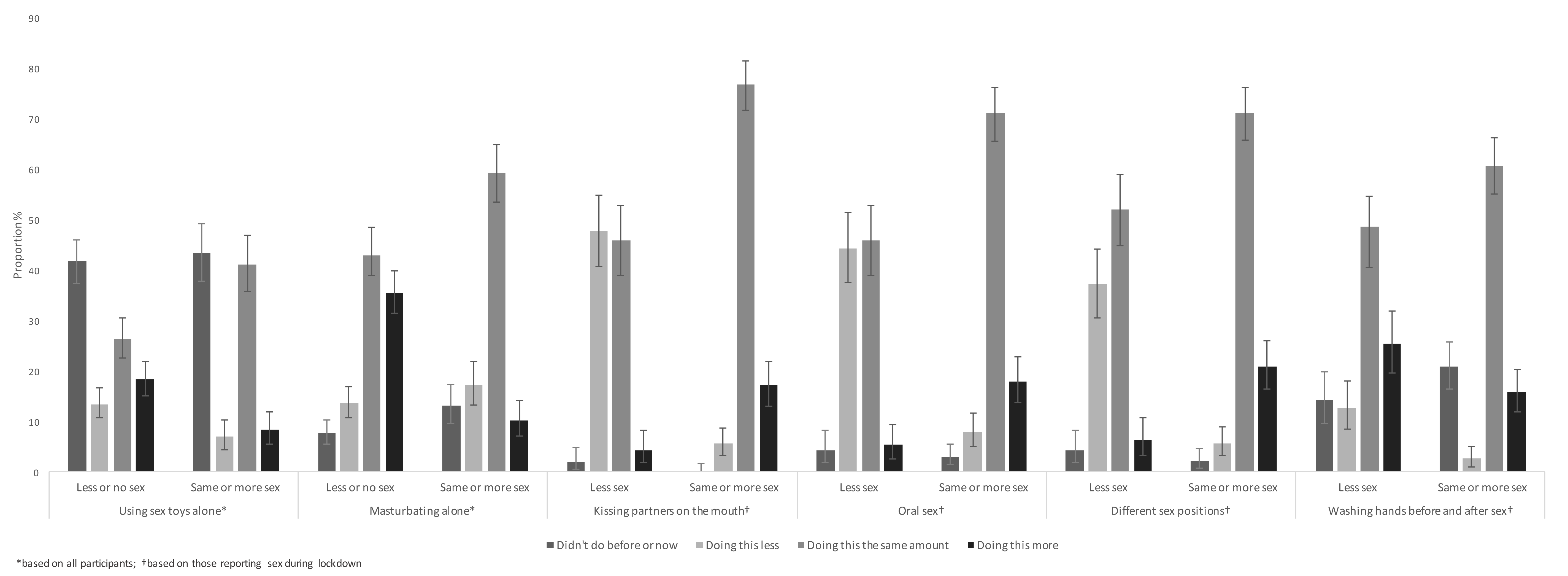
Impact of COVID-19 on frequency of sex practices stratified by those reporting less or no sex versus the same or more sex during lockdown.

Among those who reported sexual contact with another person during lockdown, 12.1% (62/511) reported kissing on the mouth, 13.0% (66/509) having oral sex, 15.1% (77/509) changing sexual positions, 19.8% (101/510) washing hands before and after sex and 2.6% (13/507) using condoms, dams or gloves more frequently than they did before COVID-19. When stratified by frequency of sex during lockdown, those who reported the same or more sex during lockdown were more likely to report kissing (17.2% vs 4.4%; diff=12.8%; 95%CI: 7.7, 17.9), oral sex (17.9% vs 5.4%; diff=12.6%; 95%CI: 7.3, 17.9) and changing sexual positions (20.9% vs 6.4%; diff=14.5%; 95%CI: 8.8, 20.2) during COVID-19 than those who reported less or no sex. However, they were less like to report washing hands before and after sex (15.8% vs 25.5%; diff=-9.6%; 95%CI: -16.9, -2.4, Figure 1).

### Dating app use

Overall 42.1% (406/965) participants reported using dating apps in 2019 with a median of two apps each (IQR=1-2; range: 1-7). The most popular apps were Tinder (78.7%), Bumble (64.8%), Hinge (15.7%), and Grindr (11.7%). App use in 2019 was reported most among MSM (61/91; 67.0%), WSW (107/201; 53.2%) and among singles (220/329; 66.9%) and least among those in cohabitating relationships (71/346; 20.5%). During lockdown, app use was reported most among MSM (40/89; 44.9%) and singles (198/325;60.9%) and least among those in cohabitating (23/344; 7.3%) and non-cohabitating (20/245;8.2%) relationships. Overall, app use decreased significantly during lockdown (42.1% vs 27.3%; difference[diff]= -14.8%; 95%CI: -17.6, -11.9, Table 2). The decrease in app use was significantly greater for those aged 18-29 years compared with those aged 30+ (diff=-18.0% vs -8.8%, p=0.03) and for WSW compared with heterosexuals (diff -25.9 vs -10.6, p<0.01). The decrease in app use was significantly smaller for singles compared with those in cohabitating relationships (-5.9% vs - 13.3%, p<0.01). No other differences were found.

**Table 2.** Use of dating apps and online dating in lockdown compared with 2019

| Variable | Used apps in 2019<br>n/N (%) | Used apps<br>since lockdown<br>n/N (%) | Difference (95%CI) † |
| --- | --- | --- | --- |
| **Overall** | 406/965 (42.1) | 260/953 (27.3) | -14.8 (-17.6, -11.9) |
| **Gender** |  |  |  |
| Male | 117/247 (47.4) | 85/241 (35.3) | -12.1 (-17.3, -6.9) |
| Female | 270/673 (40.1) | 162/669 (24.2) | -15.9 (-19.4, -12.4) |
| Gender diverse* | 18/42 (42.9) | 13/41 (31.7) | -11.1 (-24.6, 2.3) |
| **Age** |  |  |  |
| 18-29 | 309/635 (48.7) | 193/627 (30.8) | -18.0 (-21.8, -14.1) |
| 30+ | 96/323 (29.7) | 67/320 (20.9) | -8.8 (-5.1, -12.4) |
| **Sexual identity‡** |  |  |  |
| Heterosexual | 221/632 (35.0) | 152/624 (24.3) | -10.6 (-13.9, -7.3) |
| MSM | 61/91 (67.0) | 40/89 (44.9) | -22.1 (-31.2, -13.0) |
| WSW | 107/201 (53.2) | 55/201 (27.4) | -25.9 (-33.0, -18.8) |
| Other | 15/38 (39.5) | 12/37 (32.4) | -7.0 (-22.3, 8.2) |
| **Relationship Status§** |  |  |  |
| In a relationship and cohabitating | 71/346 (20.5) | 25/344 (7.3) | -13.3 (-16.8, -9.7) |
| In a relationship but not cohabitating | 88/248 (35.5) | 20/245 (8.2) | -27.3 (-33.5, -21.1) |
| Single | 220/329 (66.9) | 198/325 (60.9) | -5.9 (-11.0, -0.1) |
| Other | 25/37 (67.6) | 17/36 (47.2) | -20.3 (-39.0, -1.7) |

\*gender diverse includes trans and non-binary; †the difference in proportions lockdown-2019; calculated from logistic regression model; ‡MSM=men who have sex with men (including bisexual men); WSW=women who have sex with women (including bisexual women); Other=asexual and non-binary, and those who selected 'something else'; § 'In a relationship' includes those who selected married/de facto/boyfriend/girlfriend/LAT (living apart together) or long distance relationship; 'cohabiting' defined as living with partner/s; Single includes those who selected single/divorced/widowed (and did not select another relationship); Other includes people who selected polyamorous or multiple partners (and did not select another relationship option, such as single), and those who indicated in comments that they were casually dating, but not exclusive.

App use during lockdown increased significantly for chatting/texting (89.8% vs 94.5%; diff=4.7%; 95%CI:1.0, 8.5) and for virtual dates (2.6% vs 17.2%; diff=14.6%; 95%CI:10.1, 19.2), and decreased significantly for face-to-face dates (69.1% vs 16.0%; diff=-53.1%; 95%CI: -59.5, -47.2) and hook-ups (46.7% vs 14.8%; diff=-31.8%; 95%CI: -37.8, -25.8, Figure 2). The change in app use between time periods did not vary by gender but varied significantly by age for swapping pictures (diff =-9.2% for 18-29 years versus 4.9% for 30+; p=0.02) and by sexuality for hook-ups (diff=-63.9% MSM versus -22.9% for heterosexuals; p<0.01). No other differences were found (Supplementary Table 3).

**Figure 2.**
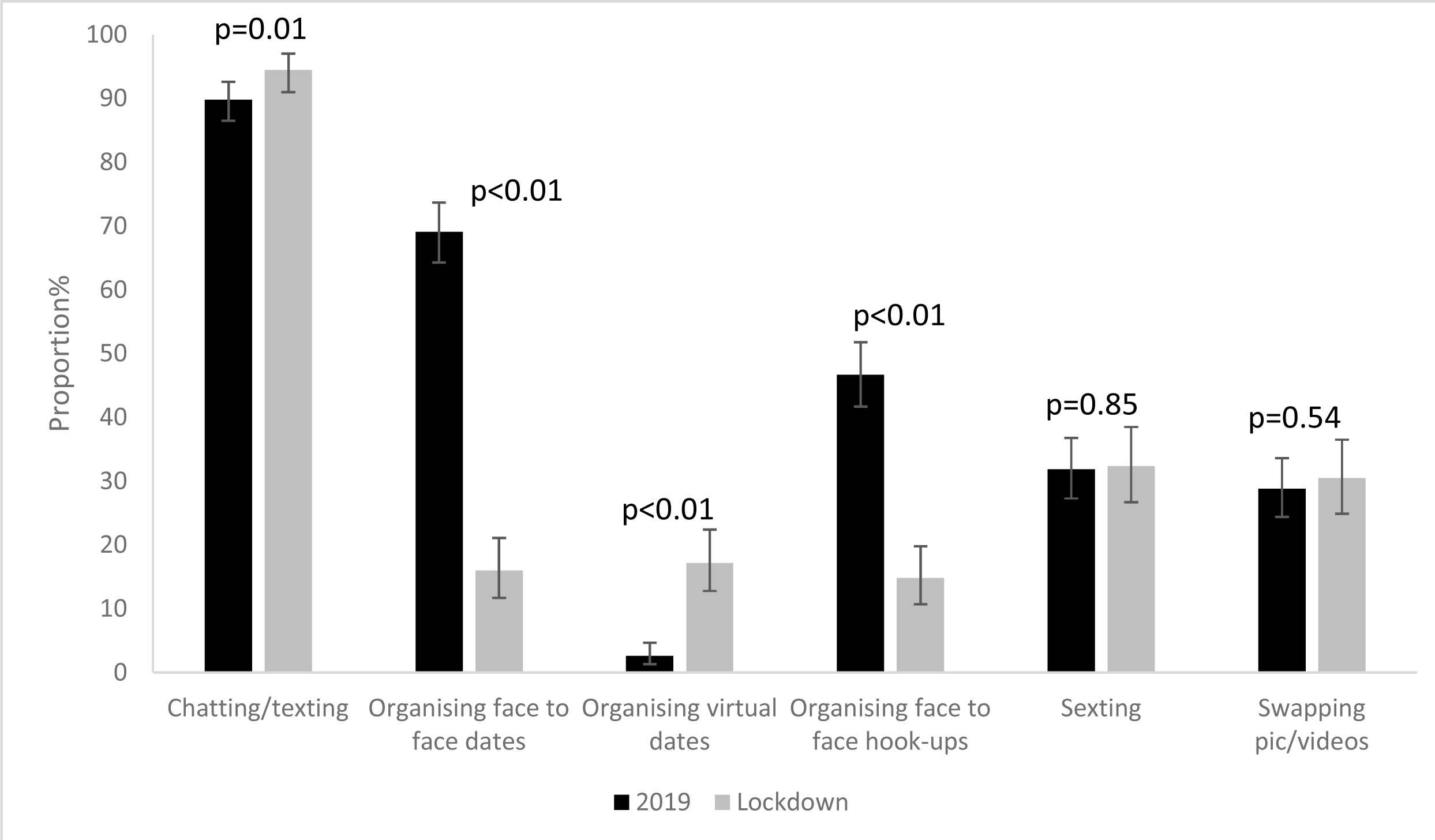
Use of dating/hook-up apps or websites in 2019 and during lockdown

### Health service use and STI diagnosis

Overall, 26.5% (211/797) reported a telehealth consultation with a GP during lockdown, 0.4% (3/797) reported accessing online STI testing and 5.4% (43/797) reported using an online pharmacy during lockdown. Few (1.2%; 10/819) reported being diagnosed with an STI during lockdown.

## DISCUSSION

Findings presented here demonstrate clear changes in sexual activity and sexual practices during the peak of the COVID-19 pandemic (to-date) in Australia and the lockdown measures it prompted. Our findings show a decline in sexual activity during the lockdown period with more than half of participants (53.5%) reporting having less sex during lockdown as compared to 2019. These patterns were most stark among those who reported being single, with 69.1% reporting less sex compared to those who reported being in a relationship. Somewhat unsurprisingly, of those reporting sexual activity during lockdown, sexual partners were most often regular partners, with few reporting sexual activity with casual hook-ups (7.8%). While research on the impact of COVID-19 on sexual behaviour is currently limited, data from a cross-sectional study conducted in the United Kingdom among those self-isolating reported similar results, with participants who were married or in a domestic relationship more likely to report sexual activity in the past week than their single counterparts[17].

Alongside declines in sexual activity, our findings also demonstrate changes in sexual practices. As could be anticipated given reduced opportunity for meeting partners, our findings show an increase in solo sex activities, including masturbation and using sex toys, particularly among those reporting less or no sex during lockdown. Indeed, adult stores in Australia reported a surge in sales during lockdown[18]. We were also interested in whether participants had changed their normal sexual practices or were engaging in additional hygiene practices, like washing their hands before and after sex, due to COVID-19. Although some participants indicated more frequently washing their hands before and after sex, over all we saw little change in partnered sex practices.

Contrary to our initial assumptions that dating app use would increase during lockdown, overall our findings showed a marked decrease in use. In particular, we saw significant declines in use among people who identified as female and aged 18-29 years. Dating apps are often used to facilitate in-person sexual and romantic connections[19], and the physical distancing enforced during lockdown perhaps drove usual users off the platform. Of those still using apps during lockdown, while some continued to organise in-person dates/hook-ups, we saw a significant increase in use for chatting/texting and organising virtual dates. Interestingly, rates of sexting or swapping intimate pictures did not significantly change between 2019 and lockdown. Further, only a small proportion of participants reported being diagnosed with STIs during lockdown and few reported accessing STI testing. However, given that some participants reported arranging in-person dates/hook-ups during lockdown, ready access to STI screening and treatment services during the COVID-19 pandemic is vital.

Our findings should be interpreted within their limitations. Namely, we used convenience sampling to recruit participants, and our resultant sample was largely homogenous, with most participants identifying as female, aged <30 years, and well educated. We also had an over-representation of WSW. Further limitations include recall bias, particularly for data on activity during 2019, and missing data for several variables (although this was usually ≤13%). Several participants also initiated the survey but did not complete it. Those who completed the survey were more likely to be older than those who did not. However, our study is novel in providing unique insight into changes in sexual activity during the peak of COVID-19 lockdown restrictions (to-date) among a cohort of people living in Australia and its strengths will be realised in subsequent cohort analyses of future waves of data.

The COVID-19 pandemic and the lockdown measures it prompted clearly impacted the sexual activity of people living in Australia. Although restrictions are beginning to ease across most of the country, recent spikes in cases in Melbourne [20], the second largest city in Australia, serve as a reminder that the pandemic is still with us and probably will be for some time. It is therefore essential to continue to monitor changes in sexual activity, and associated implications for sexual and reproductive health. In the short term, as restrictions lift and people increasingly engage in casual sex, sexual health organisations have produced guidelines for reducing risk of COVID-19 transmission during these encounters, and are encouraging regular HIV and STI screening[13]. Others warn of the continued impact of the pandemic on sexual and reproductive health, including reduced access to abortion services and an increase in intimate partner violence[21]. Whether or not Australia will experience an increase in fertility, as has been observed after high-mortality disasters like the 2004 Indonesian tsunami[22], or a long-term fertility reduction as seen in Sweden after the 1918 pandemic[23] is yet to be seen.

## Data Availability

No data are available.

## Acknowledgments

We would like to thank everyone who generously gave their time to complete our survey.

## Competing interests

EC reports grants from the National Health and Medical Research Council, outside the submitted work. JH is supported by a National Health and Medical Research Council Fellowship (1136117). The other authors report no competing interests.

## Funding

This study did not receive any external funding.

## Author contribution

All authors contributed to the design and development of the survey. JC was responsible for administering the survey. HB and JH conducted the analysis. JC, HB, JH and FK interpreted the results and drafted the manuscript. All authors contributed to the revision of draft iterations of the manuscript prior to submission.

---

**Supplementary Table 1:** Frequency of sexual activity in lockdown compared with 2019 by gender, age, sexuality and relationship status

|  | N | No sex now & no sex in 2019<br>n (%) | Less sex compared with 2019<br>n (%) | Same amount of sex as 2019<br>n (%) | More sex than in 2019<br>n (%) | p value* |
| --- | --- | --- | --- | --- | --- | --- |
| **Overall** | 883 | 77 (8.7) | 472 (53.5) | 208 (23.6) | 126 (14.3) | N/A |
| **Gender** |  |  |  |  |  |  |
| Male | 214 | 21 (9.8) | 127 (59.4) | 40 (18.7) | 26 (12.2) | 0.23 |
| Female | 629 | 51 (8.1) | 322 (51.2) | 160 (25.4) | 96 (15.3) |  |
| Gender diverse† | 39 | 5 (12.8) | 22 (56.4) | 8 (20.5) | 4 (10.3) |  |
| **Age** |  |  |  |  |  |  |
| 18-29 years | 578 | 51 (8.8) | 324 (56.1) | 111 (19.2) | 92 (15.9) | <0.01 |
| 30+ years | 300 | 25 (8.3) | 144 (48.0) | 97 (32.3) | 34 (11.3) |  |
| **Sexual identity ‡** |  |  |  |  |  |  |
| Heterosexual/straight | 572 | 53 (9.3) | 289 (50.5) | 149 (26.1) | 81 (14.2) | <0.01 |
| MSM | 80 | 5 (6.3) | 56 (70.0) | 14 (17.5) | 5 (6.3) |  |
| WSW | 195 | 10 (5.1) | 110 (56.4) | 38 (19.5) | 37 (19.0) |  |
| Other | 34 | 8 (23.5) | 16 (47.1) | 7 (20.6) | 3 (8.8) |  |
| **Relationship status§** |  |  |  |  |  |  |
| In a relationship and cohabiting | 321 | 5 (1.6) | 108 (33.6) | 146 (45.5) | 62 (19.3) | <0.01 |
| In a relationship, not cohabiting | 221 | 10 (4.5) | 129 (58.4) | 43 (19.5) | 39 (17.7) |  |
| Single | 307 | 62 (20.2) | 212 (69.1) | 12 (3.9) | 21 (6.8) |  |
| Other | 31 | 0 (0.0) | 21 (67.7) | 6 (19.4) | 4 (12.9) |  |

\* Chi Square p value; † gender diverse includes trans and non-binary; ‡ MSM=men who have sex with men (including bisexual men); WSW=women who have sex with women (including bisexual women); Other=asexual and non-binary, and those who selected 'something else'; § 'In a relationship'= married/de facto/boyfriend/girlfriend/LAT (living apart together) or long distance relationship; 'cohabiting'=living with partner/s; Single=single/divorced/widowed (and did not select another relationship); Other=polyamorous or multiple partners (and did not select another relationship option, such as single), and those who indicated in comments that they were casually dating, but not exclusive.

**Supplementary Table 2:** Type of sex partner/s in lockdown compared with 2019 by gender, age, sexuality and relationship status

|  | Spouse/partner |  |  | Girlfriend/boyfriend |  |  | Casual hook up |  |  |
| --- | --- | --- | --- | --- | --- | --- | --- | --- | --- |
|  | 2019<br>n/N* (%) | Lockdown<br>n/N* (%) | Diff %<br>(95% CI) | 2019<br>n/N* (%) | Lockdown<br>n/N* (%) | Diff %<br>(95% CI) | 2019<br>n/N* (%) | Lockdown<br>n/N* (%) | Diff %<br>(95% CI) |
| **Overall** | 290/822<br>(35.3) | 225/540<br>(41.7) | 6.4<br>(3.6, 9.2) | 371/822<br>(45.1) | 226/540<br>(41.8) | -3.3<br>(-7.0, -0.4) | 256/822<br>(31.4) | 42/540<br>(7.8) | -23.4<br>(-26.9, -19.8) |
| **Gender** |  |  |  |  |  |  |  |  |  |
| Male | 65/200<br>(32.5) | 49/109<br>(44.9) | 12.4<br>(5.3, 19.6) | 83/200<br>(41.5) | 42/109<br>(38.5) | -3.0<br>(-11.3, 5.4) | 81/200<br>(40.5) | 12/109<br>(11.0) | -29.5<br>(-37.2, -21.8) |
| Female | 207/589<br>(35.1) | 162/407<br>(39.8) | 4.6<br>(1.6, 7.7) | 274/589<br>(46.5) | 177/407<br>(43.5) | -3.0<br>(-7.3, 1.3) | 162/589<br>(27.5) | 27/407<br>(6.6) | -20.9<br>(-24.9, -16.8) |
| Gender diverse† | 18/33<br>(54.5) | 14/24<br>(58.3) | 3.8<br>(-13.7, 21.3) | 14/33<br>(42.4) | 7/24<br>(29.2) | -13.3<br>(-29.6, 3.1) | 13/33<br>(39.4) | 3/24<br>(12.5) | -26.9<br>(-47.4, -6.4) |
| **Age** |  |  |  |  |  |  |  |  |  |
| 18-29 | 111/531<br>(20.9) | 89/338<br>(26.3) | 5.4<br>(2.3, 8.6) | 305/531<br>(57.4) | 186/338<br>(55.0) | -2.4<br>(-7.3, 2.5) | 188/531<br>(35.4) | 31/338<br>(9.2) | -26.2<br>(-31.0, -21.5) |
| 30+ | 176/287<br>(61.3) | 135/200<br>(67.5) | 6.2<br>(1.3, 11.1) | 65/287<br>(22.6) | 39/200<br>(19.5) | -3.1<br>(-8.4, 2.1) | 68/287<br>(26.7) | 11/200<br>(5.5) | -18.2<br>(-23.2, -13.1) |
| **Sexual identity‡** |  |  |  |  |  |  |  |  |  |
| Heterosexual | 199/532<br>(37.4) | 158/354<br>(44.6) | 7.2<br>(3.8, 10.6) | 231/532<br>(43.4) | 139/354<br>(39.3) | -4.2<br>(-8.4, 0.6) | 129/532<br>(24.2) | 23/354<br>(6.5) | -17.8<br>(-21.8, -13.7) |
| MSM | 15/73<br>(20.6) | 7/33<br>(21.2) | 0.7<br>(-12.4, 13.8) | 29/73<br>(39.7) | 17/33<br>(52.5) | 11.8<br>(-4.9, 28.5) | 42/73<br>(57.5) | 7/33<br>(21.2) | -36.3<br>(-52.5, -20.2) |
| WSW | 58/183<br>(31.7) | 46/129<br>(35.7) | 4.0<br>(-1.4, 9.4) | 97/183<br>(53.0) | 63/129<br>(48.8) | -4.2<br>(-13.2, 5.9) | 72/183<br>(39.3) | 9/129<br>(7.0) | -32.4<br>(-40.4, -24.3) |
| Other | 18/34<br>(52.9) | 14/24<br>(58.3) | 5.4<br>(-12.2, 22.9) | 14/34<br>(41.2) | 7/24<br>(29.2) | -12.0<br>(-28.3, 4.3) | 13/34<br>(38.32) | 3/24<br>(12.5) | -25.7<br>(-46.0, -5.5) |
| **Relationship status§** |  |  |  |  |  |  |  |  |  |
| In a relationship and cohabiting | 222/327<br>(67.9) | 193/285<br>(67.7) | -0.2<br>(-2.7, 2.4) | 106/327<br>(32.4) | 95/285<br>(33.3) | 0.9<br>(-2.4, 4.2) | 35/327<br>(10.7) | 3/285<br>(1.1) | -9.7<br>(-12.9, -6.4) |
| In a relationship, not cohabiting | 37/215<br>(17.2) | 20/141<br>(14.2) | -3.0<br>(-7.9, 1.9) | 163/215<br>(75.8) | 115/141<br>(81.6) | 5.7<br>(-1.3, 12.8) | 58/215<br>(27.0) | 4/141<br>(2.8) | -24.1<br>(-30.7, -17.6) |
| Single | 20/243<br>(8.2) | 2/89<br>(2.3) | -6.0<br>(-10.3, -1.7) | 85/243<br>(35.0) | 8/89<br>(9.0) | -26.0<br>(-34.6, -17.4) | 146/243<br>(60.1) | 32/89<br>(36.0) | -24.1<br>(-35.7, -12.6) |
| Other | 10/35<br>(28.6) | 9/24<br>(37.5) | 8.9<br>(-9.0, 26.8) | 16/35<br>(45.7) | 8/24<br>(33.3) | -12.4<br>(-34.8, 10.1) | 17/35<br>(48.6) | 3/24<br>(12.5) | -36.1<br>(-59.0, -13.1) |

Diff= difference in proportions, 2019-lockdown. Calculated from logistic regression model ; \*N=number of people reporting sex during the time period and answering the question; † gender diverse includes trans and non-binary; ‡ MSM=men who have sex with men (including bisexual men), WSW=women who have sex with women (including bisexual women), Other=asexual and non-binary, and those who selected 'something else'; § 'In a relationship'= married/de facto/boyfriend/girlfriend/LAT (living apart together) or long distance relationship; 'cohabiting'=living with partner/s; Single=single/divorced/widowed (and did not select another relationship); Other=polyamorous or multiple partners (and did not select another relationship option, such as single), and those who indicated in comments that they were casually dating, but not exclusive

**Supplementary Table 3:** Use of dating apps in lockdown compared with 2019 by participant gender, age, sexual identity and relationship status

|  | Chatting/texting |  |  | Organising face-to-face dates |  |  | Organising virtual dates |  |  |
| --- | --- | --- | --- | --- | --- | --- | --- | --- | --- |
|  | 2019<br>n/N* (%) | 2020<br>n/N* (%) | Diff*<br>(95% CI) | 2019<br>n/N* (%) | 2020<br>n/N* (%) | Diff*<br>(95% CI) | 2019<br>n/N* (%) | 2020<br>n/N* (%) | Diff*<br>(95% CI) |
| **Overall** | 352/392<br>(89.8) | 242/256<br>(94.5) | 4.7<br>(1.0, 8.5) | 271/392<br>(69.1) | 41/256<br>(16.0) | -53.1<br>(-59.0, -47.2) | 10/392<br>(2.6) | 44/256<br>(17.2) | 14.6<br>(10.1, 19.2) |
| **Gender** |  |  |  |  |  |  |  |  |  |
| Male | 98/111<br>(88.3) | 77/84<br>(91.7) | 3.4<br>(-3.6, 10.4) | 63/111<br>(56.8) | 10/84<br>(11.9) | -44.9<br>(-55.0, -34.7) | 4/111<br>(3.6) | 14/84<br>(16.7) | 13.1<br>(5.1, 21.0) |
| Female | 240/264<br>(90.9) | 155/159<br>(97.5) | 6.6<br>(2.5, 10.6) | 196/264<br>(74.2) | 27/159<br>(17.0) | -57.3<br>(-64.5, -50.0) | 5/264<br>(1.9) | 26/159<br>(16.4) | 14.5<br>(8.7, 20.2) |
| Gender diverse† | 14/17<br>(82.4) | 10/13<br>(76.9) | -5.4<br>(-36.3, 25.4) | 12/17<br>(70.6) | 4/13<br>(30.8) | -39.8<br>(-78.4, -1.2) | 1/17<br>(5.9) | 4/13<br>(30.8) | 24.9<br>(1.3, 48.5) |
| **Age** |  |  |  |  |  |  |  |  |  |
| 18-29 | 274/299<br>(91.6) | 181/191<br>(94.8) | 3.1<br>(-1.2, 7.4) | 202/299<br>(67.6) | 32/191<br>(16.8) | -50.8<br>(-57.7, -44.0) | 8/299<br>(2.7) | 30/191<br>(15.7) | 13.0<br>(7.8, 18.2) |
| 30+ | 78/93<br>(83.9) | 61/65<br>(93.8) | 10.0<br>(2.3, 17.6) | 69/93<br>(74.2) | 9/65<br>(13.8) | -60.3<br>(-71.8, -48.9) | 2/93<br>(2.2) | 14/65<br>(21.5) | 19.4<br>(9.9, 28.8) |
| **Sexual identity‡** |  |  |  |  |  |  |  |  |  |
| Heterosexual | 190/212<br>(89.6) | 141/148<br>(95.3) | 5.6<br>(0.6, 10.7) | 148/212<br>(69.8) | 27/148<br>(18.2) | -51.6<br>(-59.3, -43.8) | 4/212<br>(1.9) | 21/148<br>(14.2) | 12.3<br>(6.7, 17.9) |
| MSM | 54/59<br>(91.5) | 38/40<br>(95.0) | 3.5<br>(-2.6, 9.5) | 34/59<br>(57.6) | 2/40<br>(5.0) | -52.6<br>(-66.1, -39.1) | 2/59<br>(3.4) | 5/40<br>(12.5) | 9.1<br>(-0.9, 19.1) |
| WSW | 97/107<br>(90.7) | 53/55<br>(96.4) | 5.7<br>(-1.0, 12.4) | 79/107<br>(73.8) | 8/55<br>(14.5) | -59.3<br>(-71.4, -47.2) | 3/107<br>(2.8) | 14/55<br>(25.5) | 22.7<br>(10.9, 34.4) |
| Other | 11/14<br>(78.6) | 9/12<br>(75.0) | -3.6<br>(-38.6, 31.4) | 10/14<br>(71.4) | 4/12<br>(33.3) | -38.1<br>(-79.3, 3.1) | 1/14<br>(7.1) | 4/12<br>(33.3) | 26.2<br>(0.5, 51.9) |
| **Relationship status§** |  |  |  |  |  |  |  |  |  |
| In a relationship and cohabiting | 58/69<br>(84.1) | 22/23<br>(95.7) | 11.6<br>(1.5, 21.7) | 41/69<br>(59.4) | 0/23<br>(0.0) | N/A | 2/69<br>(2.9) | 4/23<br>(17.4) | 14.5<br>(-0.6, 29.6) |
| In a relationship, not cohabiting | 76/86<br>(88.4) | 16/20<br>(80.0) | -8.4<br>(-25.3, 8.5) | 63/86<br>(73.3) | 3/20<br>(15.0) | -58.3<br>(-75.7, -40.7) | 2/86<br>(2.3) | 1/20<br>(5.0) | 2.7<br>(-7.5, 12.9) |
| Single | 198/213<br>(93.0) | 188/196<br>(96.0) | 3.0<br>(-1.3, 7.2) | 148/213<br>(69.5) | 33/196<br>(16.8) | -52.6<br>(-59.8, -45.5) | 5/213<br>(2.4) | 33/196<br>(16.8) | 14.5<br>(9.3, 19.7) |
| Other | 20/24<br>(83.3) | 16/17<br>(94.1) | 1.8<br>(-8.7, 30.2) | 19/24<br>(79.2) | 5/17<br>(29.4) | -49.7<br>(-81.1, -18.4) | 1/24<br>(4.2) | 6/17<br>(35.3) | 31.1<br>(9.3, 52.9) |

**Supplementary Table 3 continued:** Use of dating apps in lockdown compared with 2019 by participant gender, age, sexual identity and relationship status
|  | Organising face-to face-hook-ups |  |  | Sexting |  |  | Swapping pictures and videos |  |  |
| --- | --- | --- | --- | --- | --- | --- | --- | --- | --- |
|  | 2019<br>n/N* (%) | 2020<br>n/N* (%) | Diff<br>(95%CI) | 2019<br>n/N* (%) | 2020<br>n/N* (%) | Diff<br>(95%CI) | 2019<br>n/N* (%) | 2020<br>n/N* (%) | Diff<br>(95%CI) |
| **Overall** | 183/392<br>(46.7) | 38/256<br>(14.8) | -31.8<br>(-37.8, -25.9) | 125/392<br>(31.9) | 83/256<br>(32.4) | 0.5<br>(-5.1, 6.2) | 113/392<br>(28.8) | 78/256<br>(30.5) | 1.6<br>(-3.5, 6.9) |
| **Gender** |  |  |  |  |  |  |  |  |  |
| Male | 68/111<br>(61.3) | 17/84<br>(20.2) | -41.0<br>(-52.2, -29.9) | 48/111<br>(43.2) | 37/84<br>(44.0) | 0.8<br>(-10.0, 11.6) | 53/111<br>(47.7) | 37/84<br>(44.0) | -3.7<br>(-13.7, 5.7) |
| Female | 103/264<br>(39.0) | 18/159<br>(11.3) | -27.7<br>(-34.7, -20.7) | 70/264<br>(26.5) | 39/159<br>(24.5) | -2.0<br>(-8.6, 4.6) | 55/264<br>(20.8) | 36/159<br>(22.6) | 1.8<br>(-4.5, 8.1) |
| Gender diverse† | 12/17<br>(70.6) | 3/13<br>(23.1) | -47.5<br>(-80.9, -14.2) | 7/17<br>(41.2) | 7/13<br>(53.9) | 12.7<br>(-13.5, 38.9) | 5/17<br>(29.4) | 5/13<br>(38.5) | 9.0<br>(-11.3, 29.4) |
| **Age** |  |  |  |  |  |  |  |  |  |
| 18-29 | 134/299<br>(44.8) | 29/191<br>(15.2) | -29.6<br>(-36.7, -22.6) | 94/299<br>(31.4) | 62/191<br>(32.5) | 1.0<br>(-5.6, 7.7) | 73/299<br>(24.4) | 56/191<br>(29.3) | 4.9<br>(-1.2, 11.0) |
| 30+ | 49/93<br>(52.7) | 9/65<br>(13.8) | -38.8<br>(-50.2, -27.5) | 31/93<br>(33.3) | 21/65<br>(32.3) | -1.0<br>(-11.9, 9.8) | 40/93<br>(43.0) | 22/65<br>(33.8) | -9.2<br>(-19.1, 0.8) |
| **Sexual identity‡** |  |  |  |  |  |  |  |  |  |
| Heterosexual | 80/212<br>(37.7) | 22/148<br>(14.9) | -22.9<br>(-30.4, -15.3) | 51/212<br>(24.1) | 35/148<br>(23.6) | -0.4<br>(-7.2, 6.3) | 41/212<br>(19.3) | 33/148<br>(22.3) | 3.0<br>(-3.3, 9.3) |
| MSM | 48/59<br>(81.4) | 7/40<br>(17.5) | -63.9<br>(-79.1, -48.6) | 37/59<br>(62.7) | 24/40<br>(60.0) | -2.7<br>(-19.7, 14.3) | 42/59<br>(71.2) | 25/40<br>(62.5) | -8.7<br>(-21.9, 4.5) |
| WSW | 47/107<br>(43.9) | 6/55<br>(10.9) | -33.0<br>(-44.9, -21.1) | 34/107<br>(31.8) | 19/55<br>(34.5) | 2.8<br>(-9.7, 15.2) | 28/107<br>(26.2) | 17/55<br>(30.9) | 4.7<br>(-7.3, 16.8) |
| Other | 8/14<br>(57.1) | 3/12<br>(25.0) | -32.1<br>(-68.4, 4.1) | 3/14<br>(21.4) | 5/12<br>(41.7) | 20.2<br>(-7.6, 48.1) | 2/14<br>(14.3) | 3/12<br>(25.0) | 10.7<br>(-7.1, 28.5) |
| **Relationship status§** |  |  |  |  |  |  |  |  |  |
| In a relationship and cohabiting | 33/69<br>(47.8) | 2/23<br>(8.7) | -39.1<br>(-54.5, -23.7) | 21/69<br>(30.4) | 9/23<br>(39.1) | 8.7<br>(-11.1, 28.5) | 24/69<br>(34.8) | 10/23<br>(43.5) | 8.7<br>(-10.2, 27.6) |
| In a relationship, not cohabiting | 26/86<br>(30.2) | 4/20<br>(20.0) | -10.2<br>(-28.3, 7.9) | 19/86<br>(22.1) | 7/20<br>(35.0) | 12.9<br>(-8.0, 33.8) | 16/86<br>(18.6) | 4/20<br>(20.0) | 1.4<br>(-15.6, 18.4) |
| Single | 111/213<br>(52.1) | 28/196<br>(14.3) | -37.8<br>(-45.3, -30.3) | 76/213<br>(35.7) | 62/196<br>(31.6) | -4.0<br>(-10.5, 2.4) | 66/213<br>(31.0) | 61/196<br>(31.1) | 0.1<br>(-5.8, 6.1) |
| Other | 13/24<br>(54.2) | 4/17<br>(23.5) | -30.6<br>(-56.9, -4.4) | 9/24<br>(37.5) | 5/17<br>(29.4) | -8.1<br>(-33.6, 17.4) | 7/24<br>(29.2) | 3/17<br>(17.6) | -11.5<br>(-33.0, 10.0) |

Diff = difference in proportions, lockdown-2019. Calculated from logistic regression model; \*N=number of people reporting sex during the time period and answering the question; †gender diverse includes trans and non-binary; ‡MSM=men who have sex with men (including bisexual men); WSW=women who have sex with women (including bisexual women); Other=asexual and non-binary, and those who selected 'something else' §'In a relationship'= married/de facto/boyfriend/girlfriend/LAT (living apart together) or long distance relationship; 'cohabiting'=living with partner/s; Single=single/divorced/widowed (and did not select another relationship); Other=polyamorous or multiple partners (and did not select another relationship option, such as single), and those who indicated in comments that they were casually dating, but not exclusive.

STROBE Statement—Checklist of items that should be included in reports of ***cross-sectional studies***

|  | Item No | Recommendation | Reported on page # |
| --- | --- | --- | --- |
| Title and abstract | 1 | (a) Indicate the study’s design with a commonly used term in the title or the abstract | 1 |
|  |  | (b) Provide in the abstract an informative and balanced summary of what was done and what was found | 2 |
| Introduction |  |  |  |
| Background/rationale | 2 | Explain the scientific background and rationale for the investigation being reported | 3 |
| Objectives | 3 | State specific objectives, including any prespecified hypotheses | 3 |
| Methods |  |  |  |
| Study design | 4 | Present key elements of study design early in the paper | 4 |
| Setting | 5 | Describe the setting, locations, and relevant dates, including periods of recruitment, exposure, follow-up, and data collection | 4 |
| Participants | 6 | (a) Give the eligibility criteria, and the sources and methods of selection of participants | 4 |
| Variables | 7 | Clearly define all outcomes, exposures, predictors, potential confounders, and effect modifiers. Give diagnostic criteria, if applicable | 4-5 |
| Data sources/ measurement | 8* | For each variable of interest, give sources of data and details of methods of assessment (measurement). Describe comparability of assessment methods if there is more than one group | 4-5 |
| Bias | 9 | Describe any efforts to address potential sources of bias | 5 |
| Study size | 10 | Explain how the study size was arrived at | 5 |
| Quantitative variables | 11 | Explain how quantitative variables were handled in the analyses. If applicable, describe which groupings were chosen and why | 5 |
| Statistical methods | 12 | (a) Describe all statistical methods, including those used to control for confounding | 4-5 |
|  |  | (b) Describe any methods used to examine subgroups and interactions | 4-5 |
|  |  | (c) Explain how missing data were addressed | 5 |
|  |  | (d) If applicable, describe analytical methods taking account of sampling strategy | N/A |
|  |  | (e) Describe any sensitivity analyses | N/A |
| Results |  |  |  |
| Participants | 13* | (a) Report numbers of individuals at each stage of study—eg numbers potentially eligible, examined for eligibility, confirmed eligible, included in the study, completing follow-up, and analysed | 5 |
|  |  | (b) Give reasons for non-participation at each stage | N/A |
|  |  | (c) Consider use of a flow diagram | N/A |
| Descriptive data | 14* | (a) Give characteristics of study participants (eg demographic, clinical, social) and information on exposures and potential confounders | 5 |
|  |  | (b) Indicate number of participants with missing data for each variable of interest | 5-10 |
| Outcome data | 15* | Report numbers of outcome events or summary measures | 5-10 |
| Main results | 16 | (a) Give unadjusted estimates and, if applicable, confounder-adjusted estimates and their precision (eg, 95% confidence interval). Make clear which confounders were adjusted for and why they were included | N/A |
|  |  | (b) Report category boundaries when continuous variables were categorized | N/A |
|  |  | (c) If relevant, consider translating estimates of relative risk into absolute risk for a meaningful time period | N/A |
| Other analyses | 17 | Report other analyses done—eg analyses of subgroups and interactions, and sensitivity analyses | 5-10 |
| **Discussion** |  |  |  |
| Key results | 18 | Summarise key results with reference to study objectives | 10 |
| Limitations | 19 | Discuss limitations of the study, taking into account sources of potential bias or imprecision. Discuss both direction and magnitude of any potential bias | 11 |
| Interpretation | 20 | Give a cautious overall interpretation of results considering objectives, limitations, multiplicity of analyses, results from similar studies, and other relevant evidence | 10-11 |
| Generalisability | 21 | Discuss the generalisability (external validity) of the study results | 10-11 |
| **Other information** |  |  |  |
| Funding | 22 | Give the source of funding and the role of the funders for the present study and, if applicable, for the original study on which the present article is based | 11 |

\*Give information separately for exposed and unexposed groups.
**Note:** An Explanation and Elaboration article discusses each checklist item and gives methodological background and published examples of transparent reporting. The STROBE checklist is best used in conjunction with this article (freely available on the Web sites of PLoS Medicine at http://www.plosmedicine.org/, Annals of Internal Medicine at http://www.annals.org/, and Epidemiology at http://www.epidem.com/). Information on the STROBE Initiative is available at www.strobe-statement.org.

## Footnotes

1 At the time of writing, Melbourne (the capital city of the state of Victoria and the second largest city in Australia) is experiencing an outbreak of COVID-19 cases. Restrictions in areas of the city have reverted to strict lockdown restrictions as of 1^st^ July 2020 and will continue for four weeks. All other states and territories continue to report no, or single digit figures of new cases each day.

## Notes

### Author Declarations

This study was approved by the University of Melbourne Human Research Ethics Committee (ID: 2056693)

